# Dynamic changes in methadone distribution for opioid use disorder treatment from 2019 to 2023

**DOI:** 10.1101/2024.11.10.24317059

**Authors:** Amelia G Golden, Brian J Piper

## Abstract

**Background:** Opioid-related overdoses are a major concern in the United States (US) stemming from the high rates of Opioid Use Disorder (OUD). The rates of US overdoses continue to exceed one-hundred thousand per year. Methadone, a medication used as a treatment for OUD, is effective in reducing cravings and withdrawal symptoms which improves the quality of life for OUD. This study analyzed the distribution of methadone to Opioid Treatment Programs (OTP) across the US including the District of Columbia and Puerto Rico.

**Methods:** The weight of methadone (in grams) was gathered from the Drug Enforcement Administration’s Automated Reports and Consolidated Ordering System (ARCOS) database and population data from the US Census Bureau. The number of OTPs per state from the ARCOS Database was obtained. GraphPad Prism Version10.2.3 was used to make figures and perform data analysis. A paired t-test compared distribution in 2019 to 2023. Datawrapper was used to create heat maps.

**Results:** Analyzing the population-adjusted percent change by state from 2019 to 2023 showed 32 states had a positive percent change (61.5%), 19 states had a negative one (36.5%), and one state had no change (1.9%). The percent change between 2019 and 2023 (+6.2%) was statistically significant (p=0.047). When looking at the number of OTPs (population-adjusted), there was a twenty-fold state-level difference in 2023.

**Discussion:** Although there was a modest increase overall, there were pronounced state-level differences in methadone distribution across the nation. An emphasis on establishing policies to make a positive change widespread over the nation is crucial. New policies are urgently needed to increase the availability of OUD treatments more widely by initiating more OTPs and a more treatment retention focused program to reverse the escalation in overdose deaths.

## Introduction

Accidental poisoning by opioid-related overdoses is a leading cause of accidental death [1]. Of the 227,039 accidental deaths in 2022 [2], over one-third were opioid-related overdoses [3]. The estimated number of overdose deaths in 2023 from synthetic opioids was 74,702 [4]. The United States (US) Food and Drug Administration (FDA) has an approved list of medications as treatment for Opioid Use Disorder (OUD) including methadone. This medication for OUD (MOUD) is a long-acting Schedule II synthetic opiate agonist administered through Opioid Treatment Programs (OTP) [5]. The administration guidelines have changed since the COVID-19 pandemic. Pre-COVID, methadone administration in an OTP was completed for 90 days before accessing take-homes and methadone was taken concurrently with counseling, increasing treatment retention [6]. Due to the inability to come into the OTP for treatments, methadone take-home doses were increased in availability during the pandemic. Stable patients were granted a 28-day supply while unstable patients received a 14-day supply [6]. Additionally, drug screening requirements were relaxed, telehealth visits opened, and counseling became optional [6].

The results from this change have not been determined definitively although information about methadone involved mortality has been contradictory [7-9]. Knowing whether the relaxed regulations for acquiring take-home dosages affected OUD treatment distribution would help to answer the question of whether to further adjust the treatment guidelines and if so, to what extent [6].

This report aims at adding to the research narrative on methadone as OUD treatment in the US since the COVID-19 pandemic [7]. Closing the gaps in knowledge on the impact of COVID-19 on OUD was the goal of this report. Using data from the comprehensive Automated Reporting and Consolidated Ordering System (ARCOS) database, the change in distribution of methadone from certified OTPs was determined. This report also determined if there were state-wide differences across the US in the patterns of methadone distribution.

## Materials and Methods

### Procedures

The distribution of weight (in grams) from ARCOS yearly drug summary database of methadone to OTP. ARCOS distinguishes methadone being used as a MOUD from being used for pain management. The grams per state, including District of Columbia (DC) and Puerto Rico (PR), were gathered from 2019 to 2023. The number of OTPs from each state were obtained from ARCOS. Estimations of state populations were collected from the US Census Bureau’s Annual Estimates of the Resident Population.

### Data-analysis

The percentage changes in the methadone distributions for OUD treatment adjusted for population were calculated between 2019 and 2020, 2020 and 2021, 2021 and 2022, 2022 and 2023, and 2019 and 2023 for each state, including DC and PR. Figures and data analysis (including t-tests where p < 0.05 was considered significant) was completed with GraphPad Prism version 10.2.3 along with Microsoft Excel. Any state with a percent change that was greater than ± 1.96 standard deviations from the mean was considered statistically significant. The fold-difference in number of OTP (highest vs lowest) was determined after excluding one state (WY) with zero OTPs. Heat maps were produced by Data Wrapper.

## Results

The US total methadone distributed to OTP from 2019 to 2013 increased. The amount went from 13.11 to 13.92 (+6.18%) metric tons. The increments with this interval showed variable changes including +0.076% from 2019 to 2020, -5.72% from 2020 to 2021, +13.10% from 2021 to 2022, and -0.50% from 2022 to 2023. However, the total number of OTPs in the US increased appreciably (+17.55%) from 1,664 in 2019 to 1,956 in 2023.

From 2019 to 2020, the overall change in distribution was not statistically significant (p=0.82). However, there were significant increases (outside the 95% CI, p < 0.05) in Ohio (+26.03%) and significant decreases in Alabama (-22.07%), New Hampshire (-24.19%), and Florida (-29.14%).

There was a major shift in distribution from 2020 to 2022. From 2020 to 2021, over two-thirds (69.2%) of states had a negative percent change. Over three-quarters (78.8%) of states had a positive percent change from 2021 to 2022. Both the change from 2020 to 2021 (p=0.0013) and 2021 to 2022 (p=0.0002) were statistically significant. From 2020 to 2021, four states had a significant decrease (p<0.05) relative to the average state: Florida (-36.08%), Nebraska (-37.61%), Kentucky (-37.77%), and New Hampshire (-37.92%). There were four states from the next year with significant increases: Kentucky (+98.36%), Nebraska (+101.17%), Florida (+123.84%), and New Hampshire (+135.29%).

However, although there was no national significance in the next year (p=0.99), there was a significant increase in Alaska (+14.31%), West Virginia (+15.46%), and North Dakota (+17.15%) and a significant decrease in District of Columbia (-15.22%).

Overall, those five years had a significant change (p=0.047). Over three-fifths (32 states, 61.5%) had a positive percent change and one-third (19, 36.5%) had a negative one. There were four states with significant changes: Ohio (+77.74%), Kentucky (+57.18%), North Dakota (+53.19%), and West Virginia (+52.67%) (Figure 1). However, the average milligrams per person of methadone distributed barely increased (+0.5%) from 2019 (52.19) to 2023 (52.45).

**Figure 1.**
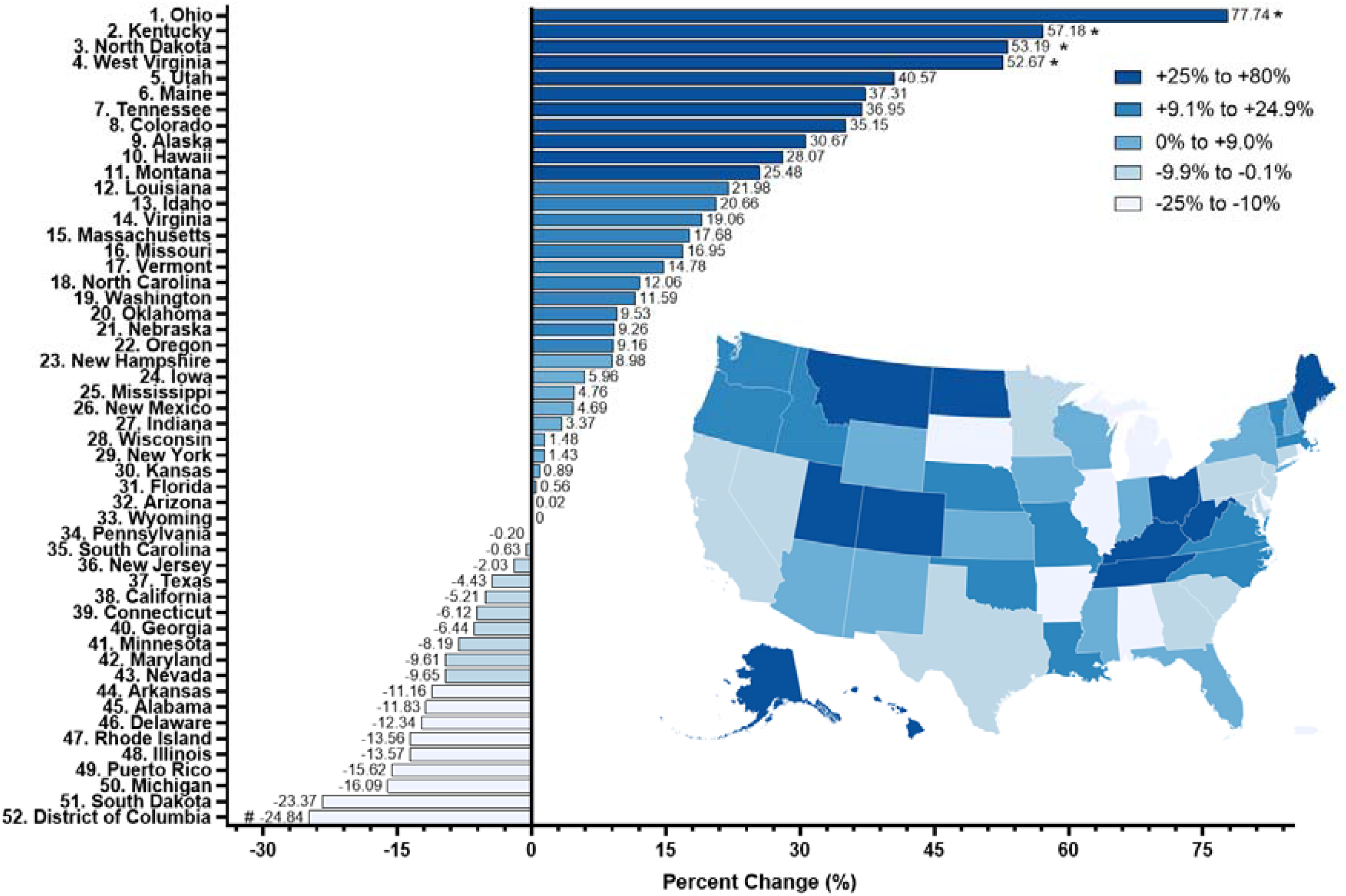
Percent change from 2019 to 2023 in methadone distribution to Opioid Treatment Programs by state (including District of Columbia and Puerto Rico) in the Drug Enforcement Administration’s Automated Reports and Consolidated Orders System. “#” indicates a state with a percent change that was between 1.5 and 1.959 Standard Deviations (SD) from the mean. “*” indicates a state considered statistically significant (p<0.05) as it had a percent change greater than ± 1.96 SDs from the mean.

The number of OTPs per million people in 2023 had a 20-fold state-level difference. There were 13 states that decreased from 2019 or 2021 to 2023. Twelve states had a decreased number of OTPs/1 million people from 2019 to 2021 but by 2023 it had reached a number greater (+23.1%) than it was in 2019 (Figure 2).

**Figure 2.**
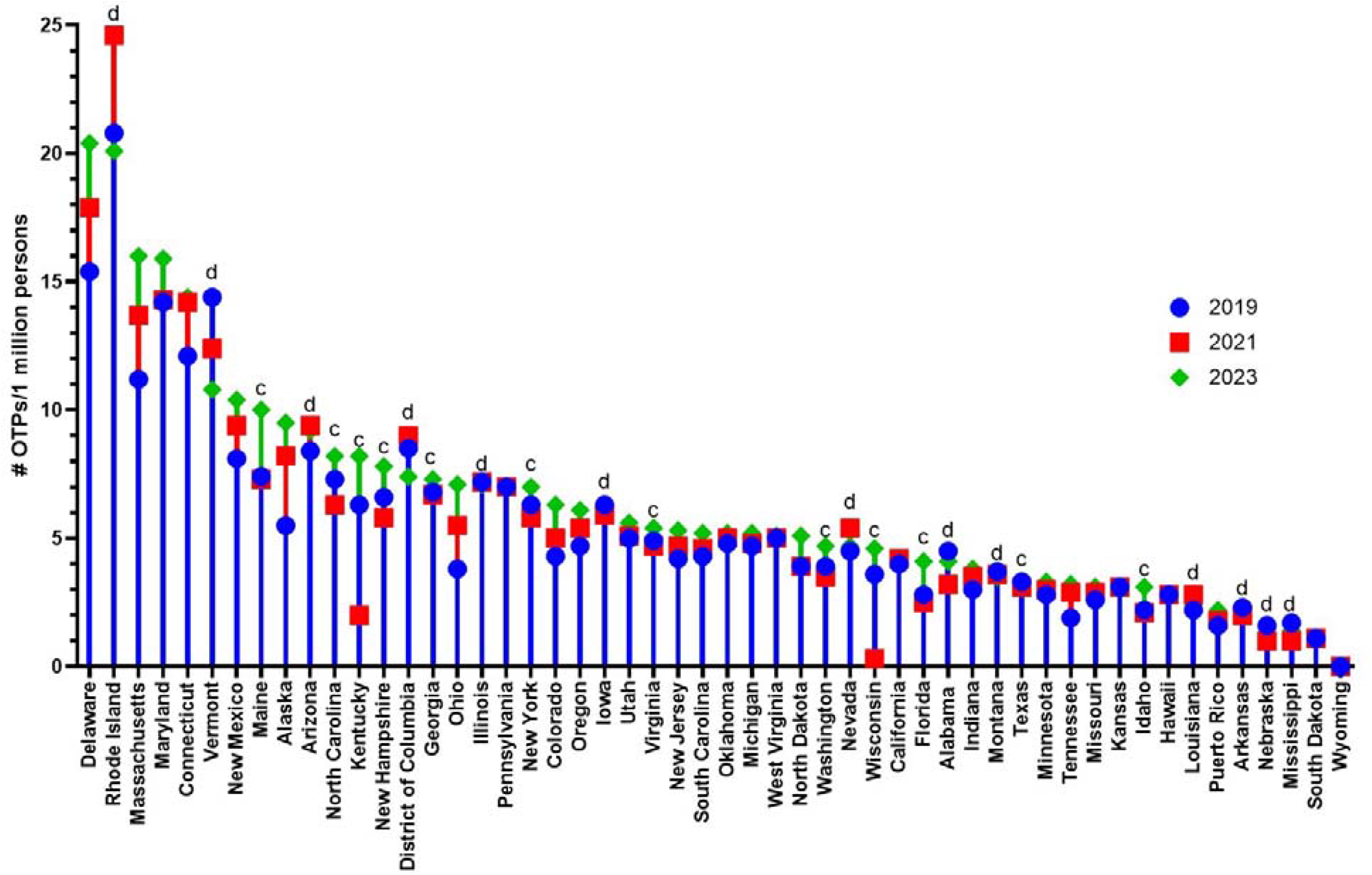
Number of Opioid Treatment Programs per one million people per state (plus District of Columbia and Puerto Rico) in 2019, 2021, and 2023 according to the Drug Enforcement Administration’s Automated Reports and Consolidated Orders System ranked in 2023. “d” indicates the thirteen states that decreased from either 2019 or 2021 to 2023. “c” indicates the twelve states that decreased from 2019 to 2021 but went back up in 2023 (2023>2019>2021).

## Discussion

There were two distinctive, interwoven novel findings in this study. First, almost two-thirds of states had an increase in methadone distributed to OTPs from 2019 to 2023. Second, most states (84.62%) had less than ten OTPs per one million people in 2023. There was an escalating rate of overdose deaths [4] even with the most states increasing in distribution. Expanding the number of OTPs, especially focusing on states with higher overdose rates but decreasing methadone availability, could help address this issue.

Examination of opioid policy of states with a positive change could assist in adapting those priorities and policies to fit more states. Ohio had a significant positive change in methadone distribution which makes it an interesting state to investigate the policies. Ohio also had less than ten OTPs per one million people but was still the top state in 2023. To maximize availability, the OTPs must be open at least six days a week. For patients to be prescribed methadone take-home dosages, there are clear guidelines [10] that must be followed and thoroughly documented by the administrator. These policies in Ohio compared to states with negative percent changes such as South Dakota, whose allusive, minimal policy has barely changed since 2017 [11], could shine light on model approaches to methadone distribution policies.

While there are strengths in using data from the Drug Enforcement Administration’s ARCOS, as they are a timely database with the ability to differentiate between the business activity of distribution which allowed for the percent change to be focused on that of just OTPs and provided the number of OTPs, there is the concern of having the weight reported instead of the number of prescriptions. This caveat does not undermine this report but opens a door for further electronic health record research to be completed identifying the profile of who is less likely to receive this evidence-based pharmacotherapy relative to others (e.g. buprenorphine) that are less efficacious in retaining patients in treatment [12].

## Conclusion

This study identified pronounced state-level disparities in methadone distribution. State disparities in number of OTPs is a prominent issue if the distribution of methadone, among other MOUDs, is to become more available. Federal or state policy adjustments could be made to maximize the beneficial effects of OTPs, particularly when methadone is combined with counseling. The unacceptably high number of overdose deaths over the past few years is an indicator of just how much work is urgently needed to permanently reverse the number of OUD fatalities.

## Data Availability

All data produced in the present study are available upon reasonable request to the authors

